# Sputum and tongue swab molecular testing for the in-home diagnosis of tuberculosis in unselected household contacts: a cost and cost-effectiveness analysis

**DOI:** 10.1101/2024.10.18.24315746

**Authors:** Charl Bezuidenhout, Lawrence Long, Brooke Nichols, Gesine Meyer-Rath, Matthew P Fox, Grant Theron, Bernard Fourie, Sharon Olifant, Adam Penn-Nicholson, Morten Ruhwald, Andrew Medina-Marino

## Abstract

**Background:** Delayed and missed diagnosis are a persistent barrier to tuberculosis control, partly driven by limitations associated with sputum collection and an unmet need for decentralized testing. Household contact investigation with point-of-care testing of non-invasive specimens like tongue swabs are hitherto undescribed and may be a cost-effective solution to enable community-based active case finding.

**Methods:** In-home, molecular point-of-care testing was conducted using sputum and tongue specimens collected from all household contacts of confirmed tuberculosis cases. A health economic assessment was executed to estimate and compare the cost and cost-effectiveness of different in-home, point-of-care testing strategies. Incremental cost effectiveness ratios of strategies utilizing different combination testing algorithms using sputum and/or tongue swab specimens were compared.

**Findings:** The total implementation cost of delivering the standard of care for a 2-year period was $84 962. Strategies integrating in-home point-of-care testing ranged between $87 844 - $93 969. The cost-per-test for in-home, POC testing of sputum was the highest at $20·08 per test. Two strategies, *Point-of-Care Sputum Testing* and *Point-of-Care Combined Sputum and Individual Tongue Swab Testing* were the most cost-effective with ICERs of $543·74 and $547·29 respectively, both below a $2,760 willingness-to-pay threshold.

**Interpretation:** An in-home, point-of-care molecular testing strategy utilizing combination testing of tongue swabs and sputum specimens would incur an additional 10.6% program cost, compared to SOC, over a 2-year period. The increased sample yield from tongue swabs combined with immediate result notification following, in-home POC testing would increase the number of new TB cases detected and linked to care by more than 800%.

**Research in context:** *Evidence before this study:* We searched PubMed for original research published between January 1, 1950 and June 30, 2024 that evaluated the cost-effectiveness of in-home POC molecular testing, as part of HCI strategies for tuberculosis. PubMed search terms used included [“household contact investigation” OR “household contact tracing”] AND “tuberculosis” AND “cost-effectiveness”. The search revealed 8 studies, of which one was removed as HCIs were leveraged for the provision of short course preventative therapy and not tuberculosis testing. None of the studies were conducted in South Africa. All seven remaining studies relied on a hub-and-spoke model of sputum collection and transportation with sputum tested at a centralized laboratory facility. Although active case finding strategies like HCIs are endorsed by the WHO to improve early case detection and treatment initiation, limited research has been done to assess its cost-effectiveness in low- and middle-income countries.

*Added value of this study:* To our knowledge, this is the first example of in-home molecular point-of-care (POC) testing as part of HCI. The use of primary data to estimate and compare the incremental cost effectiveness of different combination, in-home testing strategies utilizing alternative sample types equips policy makers with a selection of strategy options to choose from. The tradeoff between sample types with high collection yield and those with increased accuracy becomes evident in the economic analysis, highlighting the need to consider both yield and accuracy in effective clinical decision making and use-case development. The success of in-home, POC tongue swab testing of all contacts, irrespective of symptom presentation shows great promise for universal testing programs.

*Implications of all available evidence:* Results from our economic modeling provide evidence in support for the integration of in-home, POC tuberculosis (TB) testing during HCI. The use of less invasive tongue swab samples to increase sample yield in the absence of sputum expectoration highlights the value of combination testing strategies. Immediate result notification resulting from rapid, in-home POC testing shows great promise for increasing early case detection and improving treatment uptake. In-home, POC testing strategies, when incorporated into HCI could curb ongoing community transmission and reduce the overall burden of TB. Considerations for adopting novel POC testing strategies in future active case finding programs like HCI should strongly be considered.

**Summary:** We evaluated the cost-effectiveness of in-home, point-of-care TB testing of household contacts. The findings indicate that combined testing strategies using tongue swab and sputum specimens could significantly increase TB case detection, with modest additional program costs.

## INTRODUCTION

An estimated 25 000 people still die from tuberculosis (TB) every week, despite the disease being curable.^1^ Globally in 2022, 10.6 million people developed TB of which 3.1 million were never diagnosed or treated.^2^ People with missed or delayed diagnoses have elevated morbidity and mortality, drive on-going transmission, and experience increased patient and health systems costs.^3,4^ Therefore, implementation of effective, evidence-based strategies that can increase access to testing, deliver real-time point-of-care (POC) diagnosis, and reduce time to treatment initiation, are urgently required.^5^

Early screening and diagnosis is key to TB control and underpins the post-2015 END TB strategy.^4^ Active case finding strategies like HCI of known cases is widely recognized as an important component of any strategy to end TB.^5^ The WHO target product profile for new TB diagnostics place standalone, non-sputum-based near-POC tests as one of its highest priorities.^6^ For optimal POC and near-POC diagnostic tests, its cost-effectiveness is mainly a trade-off between the invasiveness of sample collection, sensitivity and specificity of the test, and the cost of resources required to conduct the test.^12^ Progress has been made towards this end. Recent studies have shown that it is feasible to integrate real-time POC molecular platforms like the GeneXpert Edge (Xpert Edge) into active case finding approaches, thus decentralizing testing services and increasing access to testing for those at-risk for TB.^7,8^ However, these studies have continued to rely on sputum-based testing alone.

As an additional sample to sputum, work to optimize the sensitivity of non-invasive tongue swab specimens when tested with rapid molecular diagnostic platforms continues to show great promise.^14^ However, despite tongue swab testing potentially having similar or diminished sensitivity when compared to sputum, the TB Home Study recently found tongue swab testing to drastically improve the number of household contacts (HHCs) of TB patients tested and increased the diagnostic yield of cases detected when integrated into HCI.^9^

Successful implementation of HCI depends on costs and affordability, while consideration for adoption is influenced by the anticipated increase in clinical effectiveness. As part of the TB Home Study, we collected resource use and prices to conduct a cost analysis of HCI from the provider perspective, comparing different testing strategies. In addition, we used decision analytic modeling of different diagnostic pathways to analyze and compare the cost-effectiveness of five novel POC testing strategies implementable as part of HCIs. Together these findings can be used to estimate the total cost and outcomes of different active case finding test strategies.

## METHODS

### Study design and participants

We used data from the TB Home Study conducted between July, 2021 and June, 2023 in the Buffalo City Metro Health District, Eastern Cape Province, South Africa. The study sought to evaluate the predictive value of pooled individual tongue swab specimens vs. sputum as a household-level triage testing strategy for TB during HCIs using the Xpert MTB/RIF Ultra cartridge (Cepheid, Sunnyvale, CA) (ie, Xpert Ultra) with the GeneXpert Edge device (Xpert Edge) as a near-POC diagnostic platform. HCIs of confirmed TB index patients were performed as described.^10^ Consenting HHCs were asked to first provide two tongue swabs and one sputum specimen. One swab from each HHC was then pooled for immediate Xpert Ultra testing in the household. The second swab was placed in Prime Store Molecular Transport Media (Longhorn Diagnostics, Inc, USA) and tested at a centralized lab. For all HHCs, irrespective of symptoms, able to produce a spot-sputum, immediate in-home testing was performed. HHCs with a positive sputum test result were immediately referred for clinic-based treatment initiation. Referral outcomes (i.e., proportion of HHCs presenting to the clinic; time-to-clinic presentation; treatment initiation outcomes) have been described elsewhere.^10^

### Cost estimation

We calculated the total implementation cost and the cost-per-test associated with each testing strategy. A provider’s perspective under routine conditions was adopted considering both fixed and variable costs reported as the testing provider. Fixed costs included building, infrastructure, and fully dedicated staff. Variable costs were based on expenditure established posteriori from quantities used, including testing supplies, consumables, Xpert Edge operating costs, and salaries for staff conducting testing. Costs were categorized and estimated separately to distinguish programmatic costs from testing costs. Programmatic costs included all expenses related to the planning and execution of HCIs. Testing costs included costs associated with HHC verification and collection, preparation, and testing of samples. Research-related expenses were excluded.

To estimate the total programmatic cost, we conducted top-down costing. Costs were categorized into equipment, personnel, laboratory costs, stationery, consumables, travel, overheads, and other miscellaneous expenses. These costs were sourced from the TB Home Study’s electronic general ledger and supplemented by interviews with key finance representatives. The total programmatic cost was estimated by summing the categories. The cost of a HCI for a single HHC was estimated by dividing the total programmatic cost by the number of HHCs reached.

Bottom-up, micro-costing was deployed to calculate the testing costs for each testing strategy. Specific inputs and their quantities needed for each testing strategy were estimated. Direct observations in combination with an electronic tracking tool built in REDCap were used to capture resources required and time needed for test activities. Estimates were used to calculate the average cost-per-test for each strategy. The total cost for each testing strategy was calculated by multiplying the cost-per-test with the expected number of tests conducted, informed by study findings.

Capital assets including furniture costs were annuitized and depreciated based on expected life-years at a 3% annual discount rate. The unit cost for running a test on Xpert Edge was calculated by dividing the cost of the platform by the total number of tests it would be able to perform across its useful lifetime. The platform can run an average of four tests per day, 892 over a year, and 4,460 over its estimated 5-year lifetime. With the total cost of the machine being $8,416, the unit cost per test would be 8,415/4,460, or US$1·89. The negotiated cost of $7·97 was used for Ultra cartridges.^17^ All cost-estimation data were obtained from the TB Home Study. All costs were inflated to 2023 prices based on annual inflation estimates provided by Statistics South Africa.^18^ Costs were then converted from South African Rand (ZAR) to US$ at the average 2023 World Bank conversion rate (1 US$ = 18·45 ZAR).^19^

### Decision analytic model design and approach

A conceptual model was developed detailing the patient diagnostic pathways for five novel HCI POC TB testing strategies and a standard-of-care (SOC) approach:

1. *SOC*: In line with South African National TB Guidelines, a HCI is conducted and all HHCs are screened using the WHO standard four-symptom screener. Those with TB-related symptoms are referred for clinic-based sputum collection and asked to return for result notification. Upon return to the clinic for result notification, those with a positive result are immediately initiated on TB treatment.^16^
2. *POC Sputum Testing*: As part of a HCI, all HHCs present are asked to expectorate sputum. Sputum samples are immediately tested using Xpert Ultra and Xpert Edge. HHCs with a positive test result are immediately referred for clinic-based treatment initiation. HHCs unable to expectorate sputum are referred to a clinic for further TB evaluation and services (i.e., TB testing; TB preventive therapy).
3. *POC Individual Tongue Swab Testing*: As part of a HCI, all HHCs present are asked to provide a tongue swab specimen for immediate, individual testing using Xpert Ultra and Xpert Edge. HHCs with a positive test result are immediately referred for clinic-based treatment initiation.
4. *POC Pooled Tongue Swabs with Confirmatory Sputum Testing*: As part of a HCI, all HHCs present are asked to provide a tongue swab specimen. A maximum of three swab specimens are pooled at a time for immediate testing in a single Xpert Ultra cartridge. Immediate, in-home confirmatory testing is done on sputum following a positive pooled test while clinic referrals are provided to those with a negative result. Confirmatory testing is done individually for each HHC able to expectorate sputum. HHCs with a positive confirmatory sputum result are immediately referred for clinic-based treatment initiation.
5. *POC Pooled Tongue Swabs with Confirmatory Tongue Swab Testing:* As part of a HCI, all HHCs present are asked to provide a tongue swab specimen. A maximum of three swab specimens are pooled at a time for immediate testing in a single Xpert Ultra cartridge. Immediate, in-home confirmatory testing is done on a tongue swab following a positive pooled test while clinic referrals are provided to those with a negative result. Confirmatory testing is done individually for each HHC able to provide a tongue swab. HHCs with a positive confirmatory result are immediately referred for clinic-based treatment initiation.
6. *POC Combined Sputum and Individual Tongue Swab Testing*: As part of a HCI, all HHCs present are asked to expectorate sputum for immediate, individual testing using Xpert Ultra and Xpert Edge. Those unable to expectorate sputum are asked to provide a tongue swab specimen for immediate, individual testing. All HHCs with a positive sputum or tongue swab result are immediately referred for clinic-based treatment initiation.

These diagnostic pathways guided the development of the final decision analytic model (Figure 1). Common elements across diagnostic pathways were identified and retained to promote consistency across activity descriptions at each decision node. All HHCs follow a similar diagnostic pathway starting with a HCI, HHC verification, Xpert Ultra testing (either in-home POC or clinic-based depending on the strategy), and treatment initiation following a positive test. The model follows HHCs through each step of the diagnostic pathway, estimating costs, transition probabilities, and outcomes.

**Figure 1:**
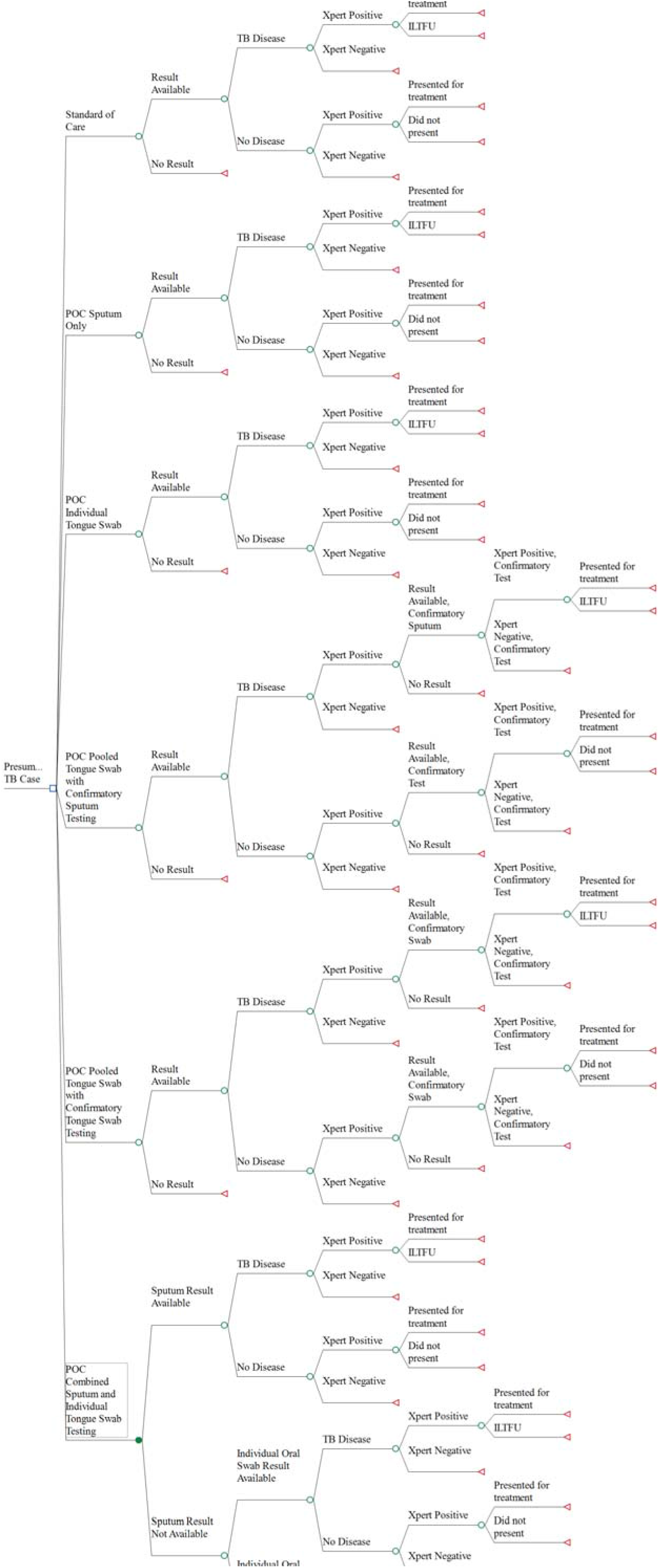
Simplified decision analytic model. The diagram shows the diagnostic pathway from testing to treatment initiation. ILTFU: Initial loss to follow up refers to HHCs who tested positive but never present for treatment initiation.

**Figure 2.**
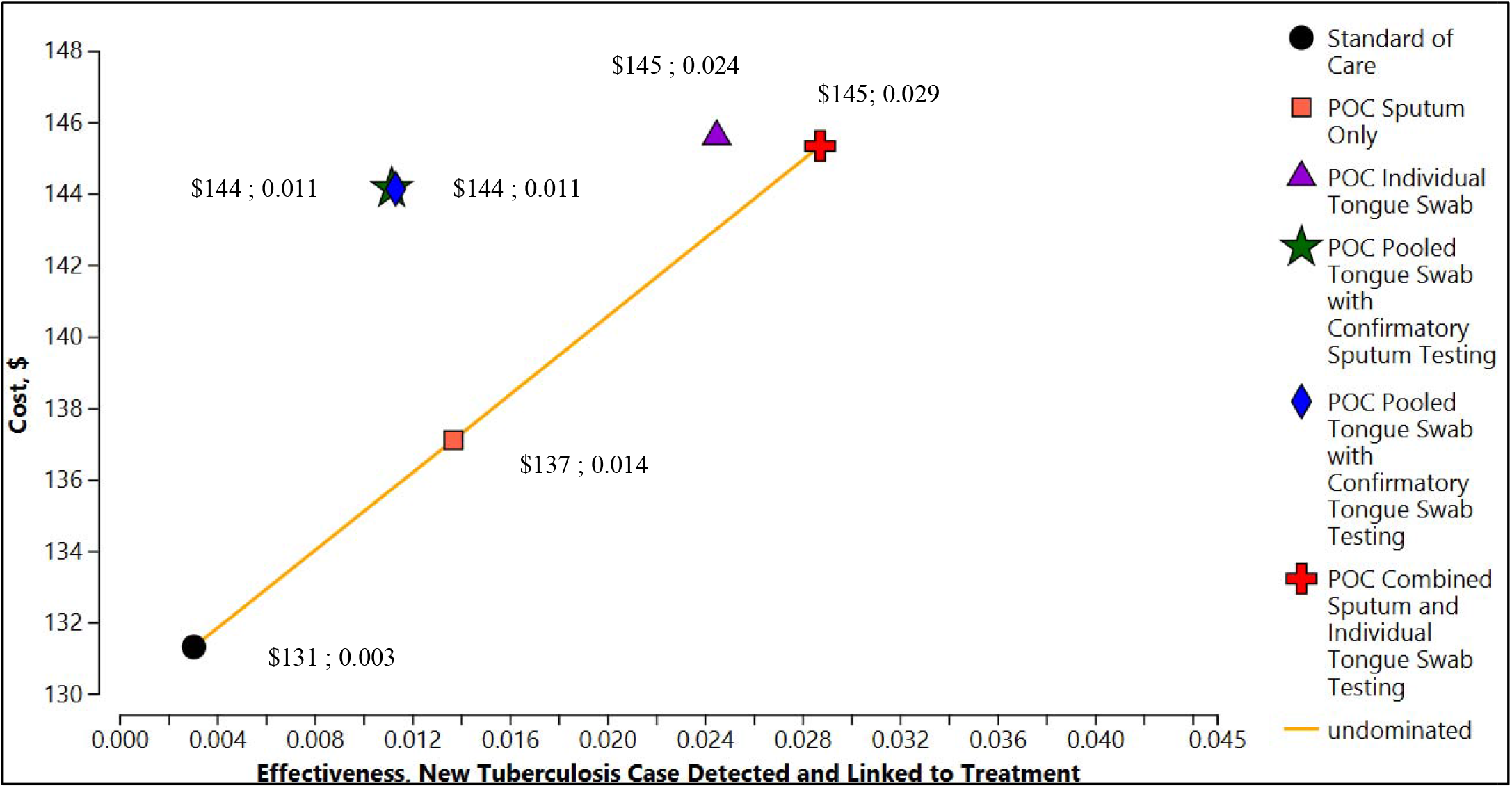
Cost-effectiveness plane of different household contact investigation testing strategies

### Outcomes and measurement of effectiveness

We derived measures of effectiveness from operational and clinical outcomes captured in the TB Home Study. Our primary cost-effectiveness outcome was the incremental cost per additional HHC with TB disease presenting at a clinic for treatment initiation, comparing each testing strategy.

### Decision analytic model for cost-effectiveness

For a testing strategy to be considered by policy makers it needs to be either as effective but less costly than SOC; or if more costly, the increase in effectiveness needs to be clinically/ diagnostically relevant enough to warrant such an increase.^12^ To this aim, incremental cost-effectiveness ratios (ICERs) were used as a metric to compare competing testing strategies.^20^ The efficacy results from the TB Home Study were used to construct a simplified decision analytic model using TreeAge Pro 2024 (TreeAge Pro, Williamston, Massachusetts, USA) to determine the cost-effectiveness of each testing strategy. The full list of parameters used in the model is provided (Supplemental Tables 1 and 2).

If the ICER for a given strategy is lower than the amount policy makers are willing to pay (WTP) for an additional unit of effectiveness, we assume it to be cost effective. We used a WTP threshold of $2,760 per HHC newly diagnosed and linked to treatment.^21^

### Sensitivity Analysis

To explore key drivers of cost and effectiveness, we performed one-way deterministic sensitivity analysis (DSA). Parameter values were changed, with the corresponding upper and lower bound values (Supplemental Table 1). The upper and lower bounds were estimated based on available literature or, in the absence of literature, as a 25% increase and decrease of the point estimate in the absence of literature. Each parameter was varied to observe its effect on the overall ICER, presented in the tornado diagram (Figure 3).

**Figure 3:**
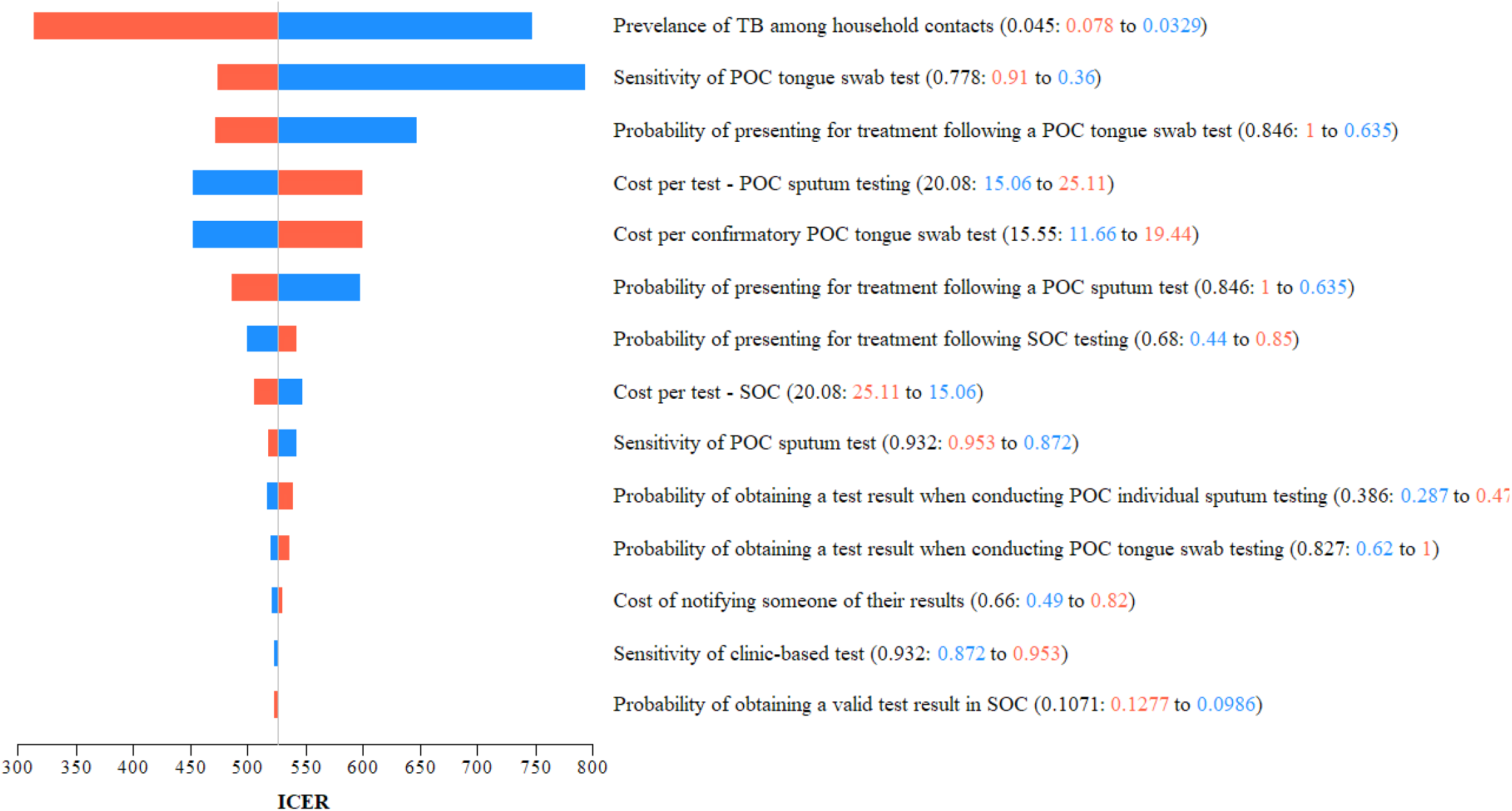
Deterministic sensitivity analysis. Tornado diagram, depicting the effect of changing individual model parameters on the ICER when comparin against POC Combined Sputum and Individual Tongue Swab Testing. The diagram depicts the base case ICER o as the difference in ICER between the two strategies to highlight the sensitivity of the ICER value to chan individual model parameters. Parameters are listed in the order of influence on cost-effectiveness. A low value indicates that cost-effectiveness has a positive correlation with the parameter value, whereas a high value (orang a negative correlation.

### Ethics approval

This study was conducted according to the ethical principles set forth in the Declaration of Helsinki, ICH-GCP, European Directive 2001/20/EC, US Code of Federal Regulations Title 21, South African Good Clinical Practice Guidelines, and other local regulatory requirements. The study protocol was approved by the University of Pretoria Human Research Ethics Committee (HREC 391/2021). Work related to the cost-effectiveness analysis was approved by Boston University Institutional Review Board (H-44118).

## RESULTS

### Cost Analysis

Supplemental Table 3 provides a summary of the total estimated implementation cost of each HCI testing strategy. The total cost of conducting HCIs (programmatic cost) was $81 327. The majority of programmatic cost (60%) was for salaries of two fully dedicated staff members followed by travel costs (24%) to transport staff to households. The total testing cost ranged from $3,635 for *SOC* to $12 642 for *POC Combined Sputum and Individual Tongue Swab Testing*. The total implementation cost ranged from $84 962 (SOC) to $93 969 for *POC Combined Sputum and Individual Tongue Swab Testing*. The integration of the Xpert Edge platform with Xpert Ultra to conduct in-home POC testing would result in a 3%-11% increase in total implementation cost, compared to SOC. The average cost-per-test (Supplemental Tables 2 and 4) was highest for sputum-based testing ($20·08). Individual tongue swab- ($19·29) and pooled tongue swab tests ($15·73) showed a 4% and 22% lower cost-per-test, respectively. The cost of Xpert Ultra was a major driver (42%) of cost-per-test.

### Probabilities

Probability data (Supplemental Table 1) for the cost-effectiveness analysis were informed by the TB Home Study and supplemented by available literature. The prevalence of TB among HHCs was estimated at 4·5% and ranged between 3·3% and 7·8%. The probability of obtaining a test result when adopting SOC was estimated to be 10·7% compared to 38·3% when in-home POC Sputum Testing was conducted. The probability of obtaining a test result from all other strategies ranged between 82·7% and 89·8%, highlighting the increased collection yield of less invasive samples. The sensitivity of different testing strategies ranged from 51·4% for *POC Pooled Tongue Swab Testing* to 93·2% for sputum-based tests. The likelihood of treatment initiation following a positive test result was 84·6% with in-home POC testing, compared to 68% for SOC.

### Cost-effectiveness analysis

Figure 2 summarizes the results of the cost-effectiveness analysis. The cost-effectiveness plane (gold line) connects undominated strategies, i.e., those not outperformed by other strategies and deemed most cost-effective. Although being the least expensive ($131·38), SOC was also the least effective. All five in-home POC testing strategies showed higher effectiveness compared to SOC albeit at a higher cost per HHC tested. *POC Pooled Tongue Swab Testing with Confirmatory Individual Tongue Swab Testing* and *POC Individual Tongue Swab Testing*, despite showing promise, were both extendedly dominated. This suggests that higher effectiveness at a lower cost could be obtained by a combination strategy located somewhere on the cost-effectiveness plane between the two undominated strategies. Table 1 summarizes the cost, effectiveness, and corresponding ICER values of the two undominated testing strategies, *POC Sputum Testing* and *POC Combined Sputum and Individual Tongue Swab Testing*. The corresponding ICERs, $543·74 and $547·29, respectively, fell below the $2 760 WTP threshold.

**Table 1.** Results of cost-effectiveness analyses of different household contact investigation testing rategies.

| Testing Strategy | Cost (\$) | Incremental Cost (\$) | Effectiveness | Incremental Effectiveness | ICER |
| --- | --- | --- | --- | --- | --- |
| Standard of Care | 131·31 |  | 0·003 |  |  |
| POC Sputum Testing | 137·10 | 5·79 | 0·014 | 0·011 | 543·74 |
| POC Combined Sputum and Individual Tongue Swab Testing | 145·33 | 8·23 | 0·029 | 0·015 | 547·29 |

### Sensitivity analysis

The DSA (Figure 3) showed prevalence of TB among HHCs, followed by the sensitivity of a tongue swab test to be the most influential parameters impacting the ICER. An increase in either estimate would result in a decrease in the ICER with clear indication that a reduction in the sensitivity of individual tongue swabs would result in a drastic increase in the ICER, albeit not exceeding the WTP threshold. None of the included parameters showed variability to the extent it would shift the ICER beyond the $2,760 WTP threshold.

## Discussion

We conducted a costing analysis to estimate the resources required to implement a HCI program in line with the South African National TB Guidelines (SOC) or when adopting one of five alternative in-home POC testing strategies. The empirical estimates derived from the TB Home Study revealed a 3%-11% increase in total implementation cost to roll out HCI with integrated in-home POC molecular testing compared to SOC. The increased cost however is due to the increase in materials and resources required to conduct additional testing, resulting in increased number of people appropriately tested for, diagnosed with, and promptly treated for TB. This finding aligns with global health priorities emphasizing the need for rapid and accurate POC diagnostics and the adoption of active case finding strategies to increase access to testing, improve patient outcomes, and reduce the number of missing TB cases.^22^

Results from the micro-costing suggest that despite being more sensitive, in-home sputum-based testing has the highest cost-per-test ($20·08). Despite lower levels of sensitivity, the 4% and 22% reduction in cost-per-test combined with increased sample yield of individual and pooled tongue swab tests could provide strong alternative solutions to increase population test coverage.^23^ The highest share (42%) of the cost-per-test is attributable to the cost of the Ultra cartridge. This finding was in line with reports from work done in other low- and middle-income settings.^11^ Despite a lower negotiated price, the cost of cartridges remain a barrier to scale-up of Ultra as a routine test. The optimal number of tongue swabs as well as optimal collection methods that yield the maximum amount of DNA remain under investigation. However, as the science evolves and the next generation of ultra-sensitive tests become available, the feasibility of using tongue swabs to address the aforementioned barriers, become more plausible.^12^

The cost per HHC when adopting *POC Sputum* and *POC Combined Sputum and Individual Tongue Swab* testing was $137 and $145, respectively. Although being $6 and $14 more expensive than SOC, these strategies would increase the likelihood of detecting TB and linking a case to treatment by 0·011 and 0·026, respectively. The associated ICERs for these two strategies were $543·47 and $547·29. These findings suggest that both *POC Sputum* and *POC Combined Sputum and Individual Tongue Swab* testing strategies were highly cost-effective given the threshold of $2,760. The increased sample yield of tongue swabs combined with immediate result notification and higher likelihood of treatment initiation following an in-home POC test, significantly improved the effectiveness of these two strategies. Findings from the cost-effectiveness analysis suggest that in a sample of 100 000 HHCs and a TB prevalence of 4·5%, a *POC Combined Sputum and Individual Tongue Swab* testing strategy would detect and link to treatment approximately 2,900 new TB cases compared to 300 (SOC).

A clinical endpoint, new TB case detected and linked to treatment, instead of a utility outcome like DALY was used to compare testing strategies. This decision was largely due to the preference of using a study-measured outcome and limiting the use of modeling assumptions. Evidence from previous economic models propose a ratio of 1:1, i.e., 1 new case detected and linked to treatment equaling to 1 DALY averted, suggesting the ICERs calculated in the current analysis would remain unchanged.^21^ The likelihood of strategies being adopted ultimately depends on the WTP of policy makers.^24^ The $2,760 threshold used in this analysis was far more conservative than the prescribed WHO-CHOICE threshold. The impact of HCI at the population level is only realized over the long term. Our current threshold is modeled off a 2-year time horizon which potentially only considers 15% of the epidemiological impact of HCI.^21^ Despite being conservative as well as potentially underestimating the long-term effects, both undominated testing strategies fell far below $2,760. These findings suggest that the adoption of in-home POC testing as part of HCI show great potential to yield positive economic returns in the long-term and should therefore be considered.

A key strength of this economic evaluation is the use of empirical cost and implementation data collected within a highly pragmatic study. This reduces reliance on modeling and findings from different contexts as both costs and effectiveness are measured in the same population.^26^ Most existing literature relies heavily on modeling analysis rather than prospectively obtained data for similar assessments.^27^ Consequently, we are confident that our results closely approximate the real-world programmatic costs of integrating POC molecular testing into HCIs. Very little research has been done to date to assesses the economic impact of implementing rapid diagnostics at POC, while none have examined its impact when integrated into HCIs.^28^

Our analysis has several limitations. Our model does not account for effects of secondary transmission, or the increased probability of transmission associated with delayed testing. Published literature on the downstream economic impact of delayed testing would suggest it’s inclusion in the current model would further strengthen the case in favor for in-home POC testing, which has shown to reduce time-to-case-notification and treatment initiation.^4,13,15^ The sputum yield parameters used in this analysis might be overestimated due to the exclusion of children, who are known to have diminished capacity to expectorate sputum.^29^

The ongoing development and refinement of rapid, POC and near-POC diagnostics holds great potential for closing gaps in the TB care cascade. However, WHO endorsement alone will not be sufficient to ensure rapid uptake and adoption into national TB programs.^30^ Integration of technologies into existing programs must be complemented by relevant system strengthening to adequately support implementation and scale-up. To promote adoption and integration, implementation studies should aim to generate the evidence needed for local policy makers to make informed decisions. This said, this study provides robust economic evidence supporting the integration of rapid POC TB testing into existing HCI strategies. Future research should aim to compare these testing strategies under more controlled conditions of a randomized control trial. Furthermore, studies should explore the scalability and sustainability of these strategies across diverse settings to inform tailored policy recommendations and optimize resource allocation in the fight against TB.

## Data Availability

All data produced in the present work are contained in the manuscript

## Author contributions

CB: data analysis, writing the manuscript

LL: writing the manuscript, supervision

BN: writing the manuscript, supervision

GMR: writing the manuscript, supervision

MPF: writing the manuscript, supervision

GT: writing the manuscript

BF: writing the manuscript

SO: writing the manuscript

APN: writing the manuscript

MR: writing the manuscript

AMM: conceptualization, funding acquisition, writing the manuscript, supervision

Data available upon request.

