## Supplementary Materials for "Sputum and tongue swab molecular testing for the in-home diagnosis of tuberculosis in unselected household contacts: a cost and cost-effectiveness analysis"

**Supplemental Materials:**

| Parameter | Base-case value | Low | High | Source |
| --- | --- | --- | --- | --- |
| Prevalence of tuberculosis among household contacts | 0∙045 | 0∙033 | 0∙078 | ^1–4^ |
| SOC |  |  |  |  |
| Probability of test result available | 0∙107 | 0∙099 | 0∙128 | ^5,*^ |
| Sensitivity of test | 0∙932 | 0∙872 | 0∙953 | ^6–8^ |
| Specificity of test | 0∙964 | 0∙963 | 0∙981 | ^6–8^ |
| Probability of presenting for treatment initiation | 0∙680 | 0∙440 | 0∙850 | ^9,10^ |
| POC Sputum Testing |  |  |  |  |
| Probability of test result available | 0∙383 | 0∙287^†^ | 0∙479^†^ | ^11,12,*^ |
| Sensitivity of test | 0∙932 | 0∙872 | 0∙953 | ^13–15^ |
| Specificity of test | 0∙964 | 0∙963 | 0∙981 | ^13–15^ |
| Probability of presenting for treatment initiation | 0∙846 | 0∙635^†^ | 1^†^ | ^*^ |
| POC Individual Oral Swab Testing |  |  |  |  |
| Probability of test result available | 0.827 | 0∙620^†^ | 1^†^ | ^*^ |
| Sensitivity of test | 0∙778 | 0∙360 | 0∙910 | ^16,17^ |
| Specificity of test | 0∙930 | 0∙660 | 1^†^ | ^16,17^ |
| Probability of presenting for treatment initiation | 0∙846 | 0∙635^†^ | 1^†^ | ^*^ |
| POC Pooled Oral Swab with Confirmatory Sputum Testing |  |  |  |  |
| Probability of pooled test result available | 0.898 | 0∙674^†^ | 1^†^ | ^*^ |
| Sensitivity of test | 0∙514 | 0∙333 | 0∙910 | ^*^ |
| Specificity of test | 1 | 0∙660 | 1 | ^*^ |
| Probability of confirmatory test result available | 0∙682 | 0∙386 | 0∙852 | ^*^ |
| Sensitivity of confirmatory sputum test | 0∙932 | 0∙872 | 0∙953 | ^13–15^ |
| Specificity of confirmatory sputum test | 0∙964 | 0∙963 | 0∙981 | ^13–15^ |
| Probability of presenting for treatment initiation following sputum test | 0∙846 | 0∙635^†^ | 1^†^ | ^*^ |
| POC Pooled Oral Swab with Confirmatory Oral Swab Testing |  |  |  |  |
| Probability of pooled test result available | 0.898 | 0∙674^†^ | 1^†^ | ^*^ |
| Sensitivity of pooled test | 0∙514 | 0∙333 | 0∙910 | ^*^ |
| Specificity of pooled test | 1 | 0∙660 | 1 | ^*^ |
| Probability of confirmatory test result available | 0.829 | 0∙622^†^ | 1^†^ | ^*^ |
| Sensitivity of confirmatory oral swab test | 0∙778 | 0∙360 | 0∙910 | ^16,17^ |
| Specificity of confirmatory oral swab test | 0∙930 | 0∙660 | 1 | ^16,17^ |
| Probability of presenting for treatment initiation following individual oral swab test | 0∙846 | 0∙635^†^ | 1^†^ | ^*^ |
| POC Combined Sputum and Individual Oral Swab Testing |  |  |  |  |
| Probability of sputum test result available | 0∙383 | 0∙287^†^ | 0∙479^†^ | ^11,12,*^ |
| Sensitivity of sputum test | 0∙932 | 0∙872 | 0∙953 | ^13–15^ |
| Specificity of sputum test | 0∙964 | 0∙963 | 0∙981 | ^13–15^ |
| Probability of presenting for treatment initiation following sputum test | 0∙846 | 0∙635^†^ | 1^†^ | ^*^ |
| Probability of individual oral swab test result available | 0.827 | 0∙620^†^ | 1^†^ | ^*^ |
| Sensitivity of individual oral swab test | 0∙778 | 0∙360 | 0∙910 | ^16,17^ |
| Specificity of individual oral swab test | 0∙930 | 0∙660 | 1 | ^16,17^ |
| Probability of presenting for treatment initiation following individual oral swab test | 0∙846 | 0∙635^†^ | 1^†^ | ^*^ |

**Table 1: Base-case value, range, and description of variables used in cost-effectiveness analyses comparing different household contact tracing testing strategies**. *Obtained from primary study data ^†^ Bound defined as a 25% decrease from point estimate. TB = tuberculosis; SOC = standard of care; POC = point of care.

***Figure 1:* Simplified diagram of decision-analytical model**

Diagram shows pathways of diagnosis and linkage to treatment initiation. ILTFU: Initial loss to follow up refers to contacts who tested positive but never present for treatment initiation.

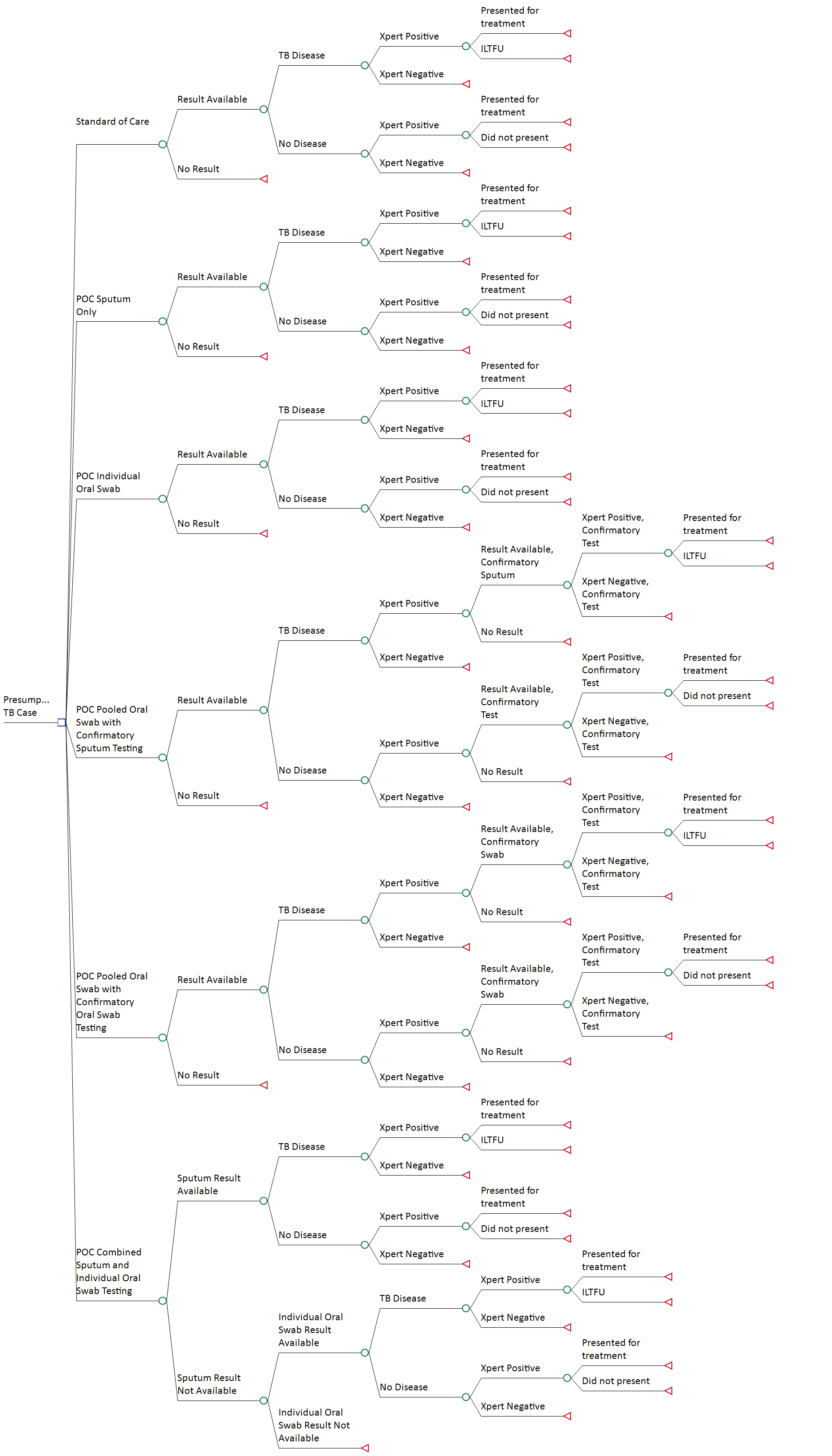

| Parameter | Base-case value ($) | Low ($) | High ($) | Source |
| --- | --- | --- | --- | --- |
| Household contact tracing cost per contact | 129∙09 | 96∙82^†^ | 161∙36^†^ | ^*^ |
| Cost of result notification | 0∙66 | 0∙49^†^ | 0∙82^†^ | ^*^ |
| SOC |  |  |  |  |
| Cost per test | 20∙08 | 15∙06^†^ | 25∙11^†^ | ^*^ |
| POC Sputum Testing |  |  |  |  |
| Cost per test | 20∙08 | 15∙06^†^ | 25∙11^†^ | ^*^ |
| POC Individual Oral Swab Testing |  |  |  |  |
| Cost per test | 19∙29 | 14∙47^†^ | 24∙12^†^ | ^*^ |
| POC Pooled Oral Swab with Confirmatory Sputum Testing |  |  |  |  |
| Cost per pooled test | 15∙73 | 11∙80^†^ | 19∙67^†^ | ^*^ |
| Cost per confirmatory sputum test | 16∙34 | 12∙23^†^ | 20∙43^†^ | ^*^ |
| POC Pooled Oral Swab with Confirmatory Oral Swab Testing |  |  |  |  |
| Cost per pooled test | 15∙73 | 11∙80^†^ | 19∙67^†^ | ^*^ |
| Cost per confirmatory oral swab test | 15∙55 | 11∙66^†^ | 19∙44^†^ | ^*^ |
| POC Combined Sputum and Individual Oral Swab Testing |  |  |  |  |
| Cost per sputum test | 20∙08 | 15∙06^†^ | 25∙11^†^ | ^*^ |
| Cost per individual oral swab test | 15∙55 | 11∙66^†^ | 19∙44^†^ | ^*^ |

**Table 2:** Base-case cost-per-test, range, and description of costs in cost-effectiveness analyses comparing different HCI testing strategies. *Obtained from primary study data. ^†^Bound defined as a 25% increase/decrease from point estimate. SOC = standard of care; POC = point-of-care.

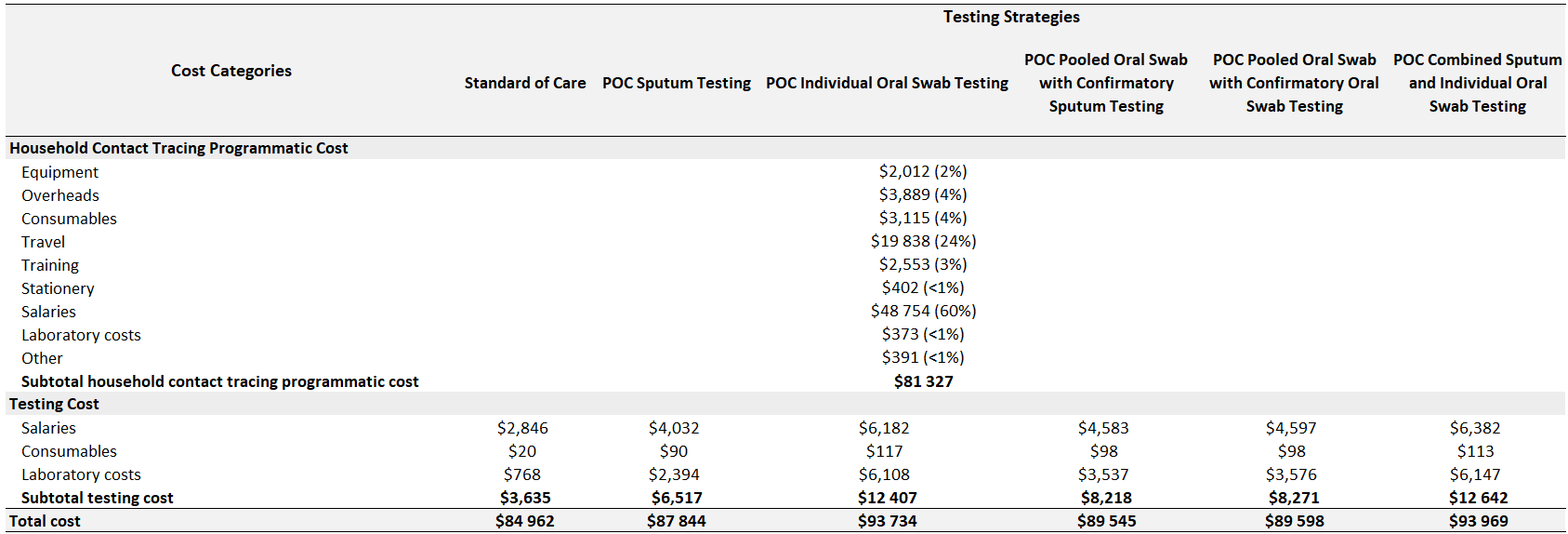

**Table 3.** Cost analysis of different household contract investigation strategies

|  | **Sputum Test** | **Individual Oral Swab Test** | **Pooled Oral Swab Test** | **Additional Sputum Test** | **Additional Oral Swab Test** |
| --- | --- | --- | --- | --- | --- |
| Salary associated with confirming HHC at the house | $3∙74 | $3∙74 | $3∙74 | - | - |
| Salary associated with testing | $6∙23 | $5∙51 | $5∙51 | $6∙23 | $5∙51 |
| Surgical gloves | 0∙13 | 0∙13 | 0∙13 | 0∙13 | 0∙13 |
| Sputum jars | 0∙13 | - | - | 0∙13 | - |
| Oral swab | - | $0∙06 | $0∙06 | - | $0∙06 |
| Ultra cartridge | $7∙97 | $7∙97 | $4∙41 | $7∙97 | $7∙97 |
| Xpert Edge machine | $1∙89 | $1∙89 | $1∙89 | $1∙89 | $1∙89 |
| **Total** | **$20∙08** | **$19∙29** | **$15∙73** | **$16∙34** | **$15∙55** |

**Table 4.** Breakdown of cost-per-test estimate. Additional Test = Additional tests include confirmatory or alternative tests conducted.

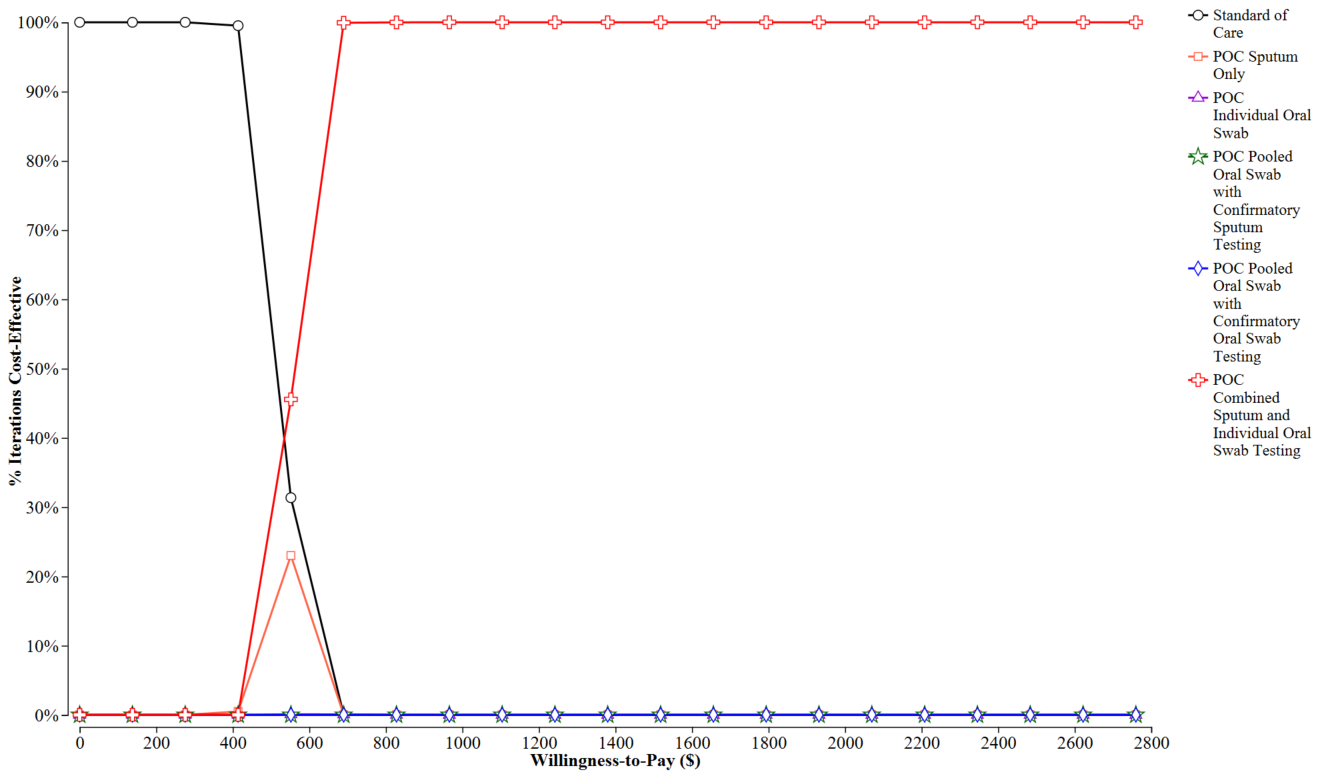

Figure 1. Cost-effectiveness acceptability curve
